# Effects of a Brief Mindfulness-Based Intervention on Pavlovian-To-Instrumental Transfer in Alcohol Use Disorder

**DOI:** 10.1101/2023.01.04.23284172

**Authors:** Annika Rosenthal, Maria Garbusow, Nina Romanczuk-Seiferth, Anne Beck

## Abstract

Pavlovian conditioned contextual cues have been suggested to modulate instrumental action and might explain maladaptive behavior such as relapse in patients suffering from alcohol use disorder (AUD). Pavlovian-to Instrumental transfer (PIT) experimentally assesses the magnitude of this context-dependent effect and studies have shown a larger PIT effect in AUD populations. Taken this into account, a reduction of the influence of cues on behavior seems warranted and one approach that could alter such cue reactivity is mindfulness. Mindfulness-based interventions have been shown to be efficient in the treatment of AUD, but underlying mechanisms are yet to be elucidated. Therefore, we aim at investigating the effect of a brief mindful body scan meditation on the magnitude of the PIT effect in AUD subjects and matched controls. Using a randomized within-subjects design, we compared the effect of a short audio guided body scan meditation against a control condition (audio of nature sounds) on PIT in healthy (n = 35) and AUD (n = 27) participants. We found no differences in PIT effect between healthy and AUD participants as well as between conditions. However, a significant interaction effect points to a decreased PIT effect after body scan meditation in AUD subjects only. These results suggest that AUD might be susceptible to mindfulness-induced changes in PIT, with these findings contributing to entangling the underlying mechanisms of the efficacy of mindfulness-based interventions in AUD.

## Introduction

Alcohol use disorder (AUD) is characterized by high relapse rates despite severe negative consequences (1). Multiple studies have indicated that alcohol-related cues can promote drug-seeking behavior in the context of AUD which has been termed cue reactivity (2). In particular, neural activation in response to alcohol cues has been associated with drinking outcomes (3-5). In the course of AUD, conditioning processes attribute incentive salience to these cues beyond the initial hedonic experience and together with drug-related neurobiological adaptations and loss of cognitive control, individuals with AUD experience craving and continue maladaptive consumption despite negative consequences (6). The association of the reinforcing effect of stimuli and instrumental behavior has been experimentally modeled in in various ways. One paradigm that has been established, is the so-called Pavlovian-to-instrumental transfer task (PIT) (7, 8). PIT tasks include an instrumental training phase aiming at linking behavioral responses to dispense of rewards. During a phase of Pavlovian conditioning, stimulus–outcome associations are established by linking formerly neutral stimuli to reward. Finally, the established instrumental behavior is assessed in the presence of the previously conditioned stimuli during Pavlovian conditioning (9). The enhancement or suppression of instrumental responses due to the value of Pavlovian conditioned stimuli is referred to as PIT effect (10). Various studies found AUD populations to be affected by an increased PIT effect, i.e. an increased impact of conditioned cues on behavior (11). In addition, the PIT effect has been associated with relapse propensity in detoxified AUD patients (12-14).

In AUD, stress is known to be an important factor that is associated with adverse disease outcomes (15, 16). It has been proposed that stress results in alterations in cognitive function as well as in a shift towards habitual decision-making (17) and might potentially emphasize automatized cue-induced behavior (18). In this context, stress has been discussed as a potential moderator of the PIT effect by strengthening the transfer of contextual cues on experimental behavior (14, 19, 20). Other studies however, did not find stress to affect the PIT effect (21, 22). One approach to reduce stress are so-called mindfulness-based interventions (MBIs). In line with this, programs such as mindfulness-based relapse prevention (MBRP) or Mindfulness-Oriented Recovery Enhancement (MORE) have been specifically tailored to addictive disorders and have shown efficacy in increasing abstinence and decreasing measures of dependence severity (23). With regards to the underlying mechanisms of the efficacy of MBIs, a body of research suggests that these interventions reduce subjective and physiological stress, increase cognitive control and decrease the effect of addiction-related cues on behavior (24). With regard to cue-reactivity, in populations with opioid abuse, evidence for an effect of mindfulness on neurophysiological reactions to drug cues have been found (25). In addition, conditioned responses to drug cues, operationalized by salivation, were reduced after an MBI (26). In this context, in AUD patients, trait mindfulness as well as mindfulness training was associated with lower reactivity as well as increased physiological recovery towards stress-primed alcohol cues (27, 28). Integrating these findings with research showing how cues affect instrumental behavior and increase relapse susceptibility, it seems to be warranted that MBIs affect the association of Pavlovian cues and instrumental behavior, such as operationalized by the PIT effect. In light of this, we wanted to investigate whether PIT effects can be modulated by mindfulness training. Therefore, participants with AUD as well as healthy controls were subjected to a brief mindful audio-guided body scan meditation before completing the transfer phase of a newly developed single-lever PIT paradigm (29). We contrasted this meditation with the same subjects passively listening to a recording of nature sounds. We specifically hypothesized that the body scan would decrease the magnitude of the PIT effect. We furthermore expected the PIT effect to be increased in AUD compared to healthy controls according to previous research (e.g. 11).

## Methods

### Participants

35 healthy controls and 27 participants with AUD were recruited via online advertisement in Berlin, Germany. A telephone screening to confirm eligibility was performed prior to assessment. Inclusion criteria were age between 18 and 70 years, sufficient German language skills and the ability to understand the study protocol and give informed consent. AUD participants had to fulfill at least two criteria for AUD according to the Diagnostic and Statistical Manual of Mental Disorders (Fifth Edition) (DSM-5) (30) with no requirement of medically supervised alcohol detoxification or request for therapeutic intervention. Exclusion criteria were positive urine drug-screening for recreational drugs as well as use of psychoactive medication and substance use disorders other than alcohol, nicotine or cannabis (mild up to two criteria). Other criteria that led to exclusion were medical history of DSM-5 bipolar disorder, psychotic disorder, schizophrenia or schizophrenic spectrum disorder, medical history of severe head trauma or other severe central nervous system disorders as well as necessity of treatment. Study participation was financially rewarded and all participants gave written informed consent. The study adhered to the Declaration of Helsinki and was approved by the local ethics committee at Charité – University Medicine, Berlin.

### Procedure

Before the experiment, participants completed diagnostic interviews as well as questionnaire assessments. The experiment was executed as a within-subjects design and testing comprised of two consecutive sessions. Participants were assigned to the intervention conditions in randomized order which consisted of participants listening to a 25 minute audio file of either nature sounds (NS) (European forest sounds, downloaded from http://stampede.it/) or a guided body scan meditation (BS) (adapted from Kabat Zin (31)), respectively. Upon completion of other cognitive tasks, the instrumental training and Pavlovian conditioning phases of the PIT paradigm were administered. Before the transfer phase and forced-choice task (i.e. query trials, where participants had to indicate which one out of two conditioned stimuli they prefer to test that the conditioning was successful), the participants were exposed to another 6 minutes in terms of a refresher of the assigned intervention condition (please see figure 1 for details of the procedure).

**Figure 1.**
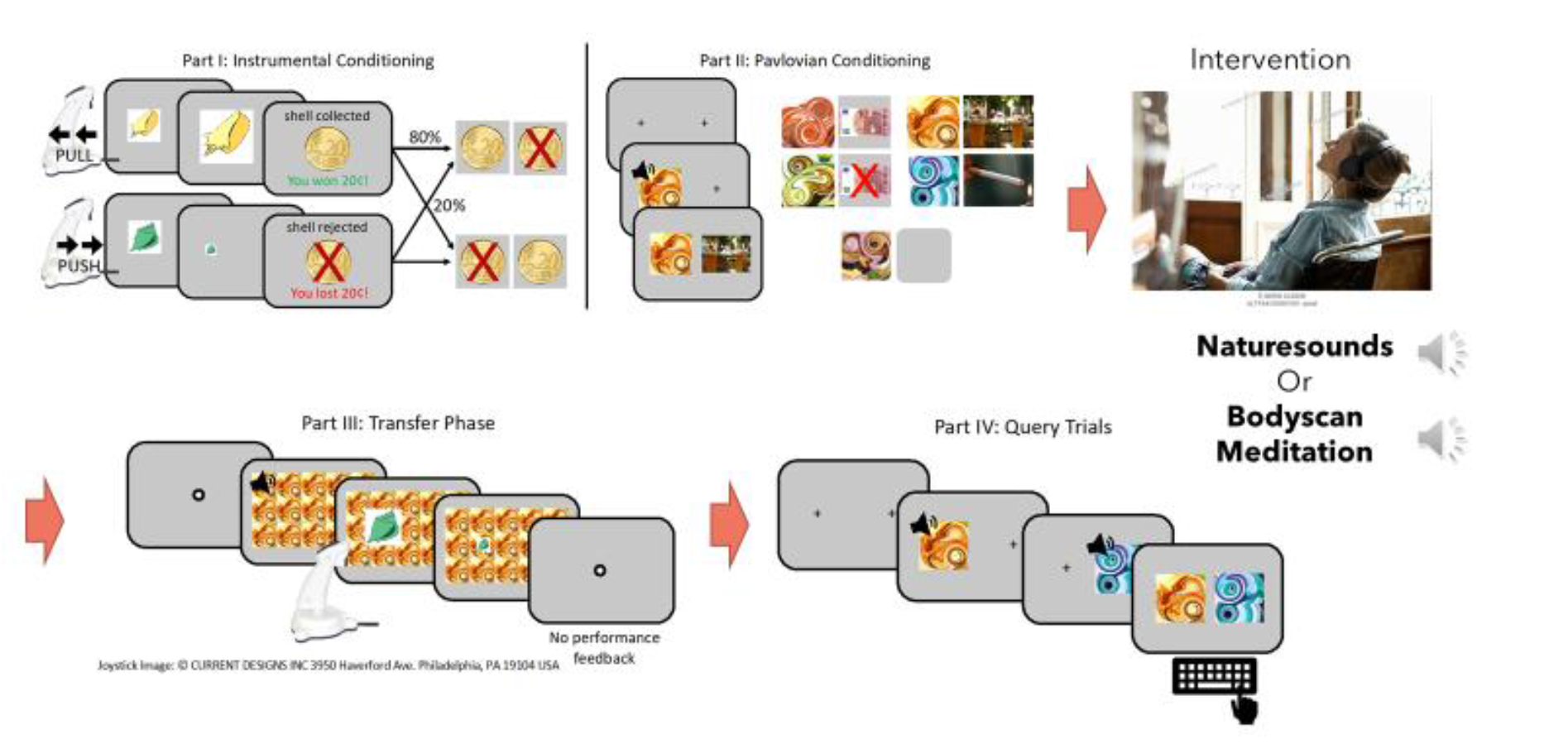
Schematic representation of the PIT procedure: In counterbalanced order, participants received a control intervention (nature sounds) or a mindfulness intervention (body scan) per experimental session. The intervention took place between Pavlovian conditioning and the actual transfer phase of the PIT task.

### PIT Paradigm

The task was programmed using Matlab 2019 (version 1.8.0_202; The MathWorks, Natick, MA) and the Psychophysics Toolbox Version 3 (PTB-3; 32). Due to the within-subjects design we used two versions that contained different stimuli in the instrumental conditioning part (shells/leaves) as well as in the Pavlovian conditioning part (different colored fractals) to prevent carry-over effects. For a detailed description of the paradigm, please see (29). The task consists of four parts (depicted in figure 1):

#### (1) Instrumental conditioning

Participants collected or rejected shells or leaves by means of a pull or push joystick movement after which they received probabilistic feedback. In the “approach trials”, collecting a shell was rewarded in 80% of the trials and penalized in 20% of the trials, and vice versa if it was rejected. For the “rejection trials”, collecting a shell was penalized in 80% of the trials and rewarded in 20% of the trials, and vice versa if it was collected. For instrumental training, a learning criterion was set to ensure that participants performed comparably in the end of instrumental training (after at least 60 trials, 80% correct decisions in 16 consecutive trials) to avoid between group effects in instrumental performance that might influence the PIT effect. Instrumental conditioning lasted a maximum of 120 trials or until the learning criterion was reached. (2) ***Pavlovian conditioning***: At the beginning of each trial, a picture of an abstract fractal accompanied by a tone (combined CS) was presented. After a 3 second delay, an unconditioned stimulus (US) (neutral: depicted by a fixation cross; positive: depicted by +10€; negative: depicted by -10€) was shown for an extra 3 seconds. Participants were instructed to pay attention to the CS-US pairs. Pavlovian conditioning ended after 80 trials. (3) ***Transfer phase***: In each trial lasting on average 3 seconds (ranging from 2-6 seconds as determined by an exponential distribution), were tiled over the background while participants were instructed to reject or collect shells/leaves according to previously learned contingencies (part 1). While no feedback was provided, participants were told that their decisions would affect the final monetary outcome. Part 3 lasted for 180 trials. (4) ***Query trials***: Over 30 trials, participants were presented with two combined CSs at a time with the instruction to choose one according to their subjective liking. All possible CS pairs were presented three times in a random order.

### Intervention conditions

During the mindfulness intervention, participants listened to a 25-minute audio file that consisted of instructions to a body scan mediation. Participants were asked to close focus on various body parts and observe any sensation without judgement while staying in the moment and letting other thoughts recede. The control condition consisted of participants’ listening to 25 minutes of nature sounds. During both conditions, participants remained seated in a comfortable chair with their eyes closed.

#### Other Measures

Next to diagnostic interviewing according to DSM-5 criteria of AUD, we administered a questionnaire commonly administered in clinical samples. The Alcohol Dependency Scale (ADS) is a measure of the severity of the participant’s dependence on alcohol and comprises of factors such as loss of control over drinking, obsessive-compulsive drinking and withdrawal symptoms (33).

To assess a proxy of premorbid intelligence, we administered a multiple choice vocabulary test, MWT-B (Mehrfachwahl-Wortschatz-Test), that is broadly used in clinical and research context in Germany (34).

Alcohol consumption was estimated by assessing quantity frequency of average consumption over the last 90 days. Participants retrospectively reported the average quantity per type of drink per occasion as well as the frequency in the given time interval in which that amount is consumed (35).

In addition, participants completed the Perceived Stress Scale (PSS) (36) widely used to measure subjectively perceived stress, as well as the Five Facets Mindfulness Questionnaire (FFMQ) (37) to assess trait mindfulness across five dimensions including nonjudging, describing, nonreacting, acting with awareness and observing.

Before and after the intervention condition, participants completed visual analog scales to assess vigilance, alertness and relaxation.

### Data analysis

Data were analyzed with MATLAB R2019b (MATLAB version 9.7, 2019; The MathWorks, Inc.) and the R System for Statistical Computing - version 4.2.2 (R Development Core Team, 2022). Demographic as well as questionnaire-based comparisons between AUD participants and HC were examined using chi-square and *t*-tests (table 1). For examination of the effect of the interventions on self-reported vigilance, alertness and relaxation, we calculated a mixed ANOVA with time (pre and post) and intervention (body scan and nature sounds) as factors.

**Table 1.** Sample characteristics of AUD and HC groups

|  | **AUD (N = 27)** | **HC (N = 35)** | **Test statistics** |
| --- | --- | --- | --- |
|  | ***M (SD)*** | ***M (SD)*** |  |
| *Demographic variables* |  |  |  |
| Age | 39.85 (11.88) | 38.51 (14.26) | $t = -0.39, p = 0.70$ |
| Sex (% female) | 17% | 59% | $\chi^2 = 11.8, p < 0.001$ |
| AUD criteria | 4.36 (2.30) | 0.06 (0.24) | $t = -10.59, p < 0.001$ |
| *Questionnaires* |  |  |  |
| <u>ADS</u> | 8.58 (4.04) | 2.60 (2.39) | $t = 6.48, p < 0.001$ |
| <u>Average Consumption per day</u> | 2.89 (2.34) | 0.54 (0.83) | $t = 4.98, p < 0.001$ |
| <u>FFMQ</u> | 136.67 (18.53) | 135.35 (17.07) | $t = 0.279, p = 0.78$ |
| <u>PSS</u> | 17.42 (6.83) | 13.97 (5.13) | $t = 2.20, p = 0.032$ |
| MWT-B | 26.50 (10.60) | 31.09 (3.53) | $t = -2.04, p = 0.051$ |

AUD = Alcohol use disorder; HC =; M = mean; SD = standard deviation; ADS = Alcohol Dependence Scale; FFMQ = Five Facets Mindfulness Scale; PSS = Perceived Stress Scale; MWT-B = Mehrfachwahl-Wortschatz-Test

To analyze the motivational component of instrumental behavior during the transfer phase, the peak velocity (in degrees per second) of the collection and rejection movements of the joystick were examined. Here, the PIT effect is reflected by the interaction of the background stimulus’ value and the peak velocity of the instrumental action. The aim of study was to assess the effect of the intervention condition as well as of group (AUD versus HC) on the PIT effect. Since we used a within-subjects design, we included session in the model to account for carry-over effects from session 1 to session 2, although administration of the intervention was done in randomized fashion.

A linear mixed-effect model (LMM) (R-package: lme4 (38)) was constructed to include the following fixed effects and their interactions: Pavlovian CS (dummy coded with 10€ as a reference), intervention condition (nature sounds or body scan, coded as 0.5 and -0.5 respectively), session (second and first session, coded as 0.5 and -0.5 respectively), group (AUD and HC, coded as 0.5 and -0.5 respectively) and instrumental response (reject and collect, coded as 0.5 and -0.5 respectively). For random effects, we included the effect of instrumental response as well as Pavlovian CS per subject as likelihood ratio tests (LRT) of random effects indicated the variance parameter of instrumental response (LRT = 2231; p < .001) and Pavlovian CS (LRT = 734; p< .001) to be significantly different from zero.

In addition, we carried out a mediation analysis testing for an association of FFMQ, PSS and ADS (please see supplementary material for details).

## Results

The groups did not differ in age, premorbid intelligence as well as trait mindfulness measured by MWT-B and FFMQ respectively. Compared to the HC group, AUD participants displayed significantly higher ADS scores, average daily consumption, perceived stress (PSS) as well as AUD criteria (please see figure 2 for distribution of AUD criteria). Finally, the AUD group contained more male participants than the HC group. All results are shown in table 1.

**Figure 2.**
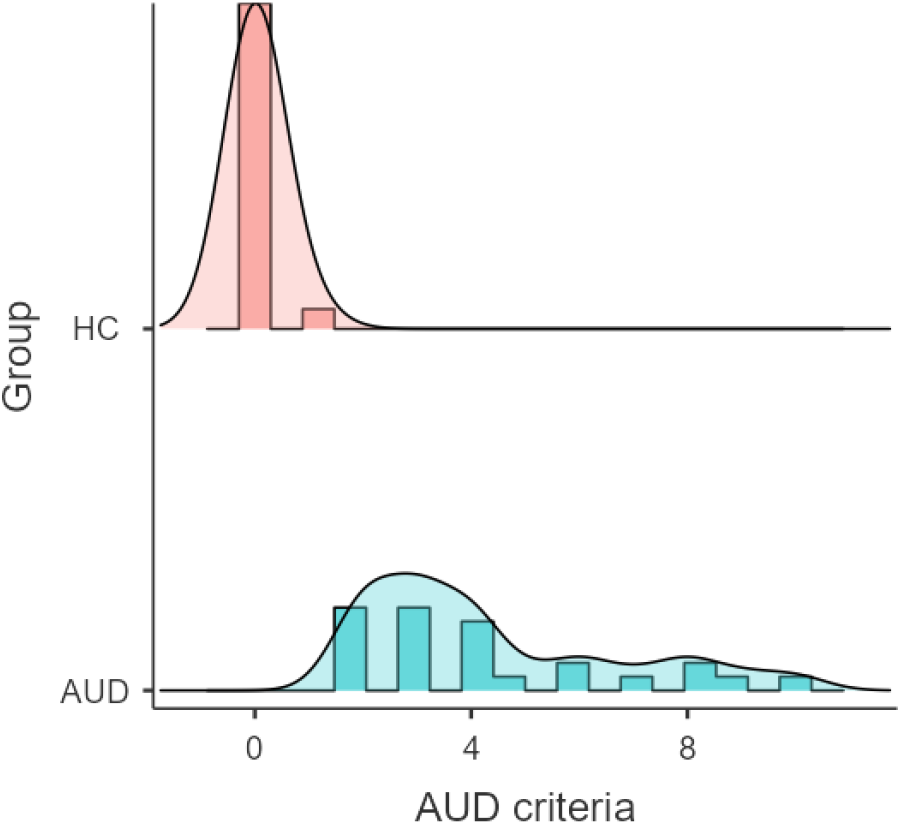
Density plot of the distribution of AUD criteria in the HC and AUD group.

The subjects’ performance during query trials indicated that there was no difference between session and conditions. The results are reported in the supplementary materials (supplementary table 1).

Regarding the transfer phase, prior to inspection of the fixed effects, we performed and compared several linear mixed models. We found the full model that contained session, group and condition to best fit our data (see table 2 for details). In extension of this, omnibus F-tests of the fixed effects are reported in supplementary table 2. Here, all four-way interactions between fixed effects were not significantly contributing to the model (ranging from p = 0.07 – p = 0.95) and were therefore omitted.

**Table 2.** Model comparisons between the full model and models that excluded fixed effects

| Model | AIC | Log Likelihood |
| --- | --- | --- |
| **1. Full model** | **176728** | **-88271** |
| 2. No session | 176914 | -88422 |
| 3. No condition | 176868 | -88399 |
| 4. No group | 176848 | -88389 |

AIC = Akaike information criterion

**Table 3.**
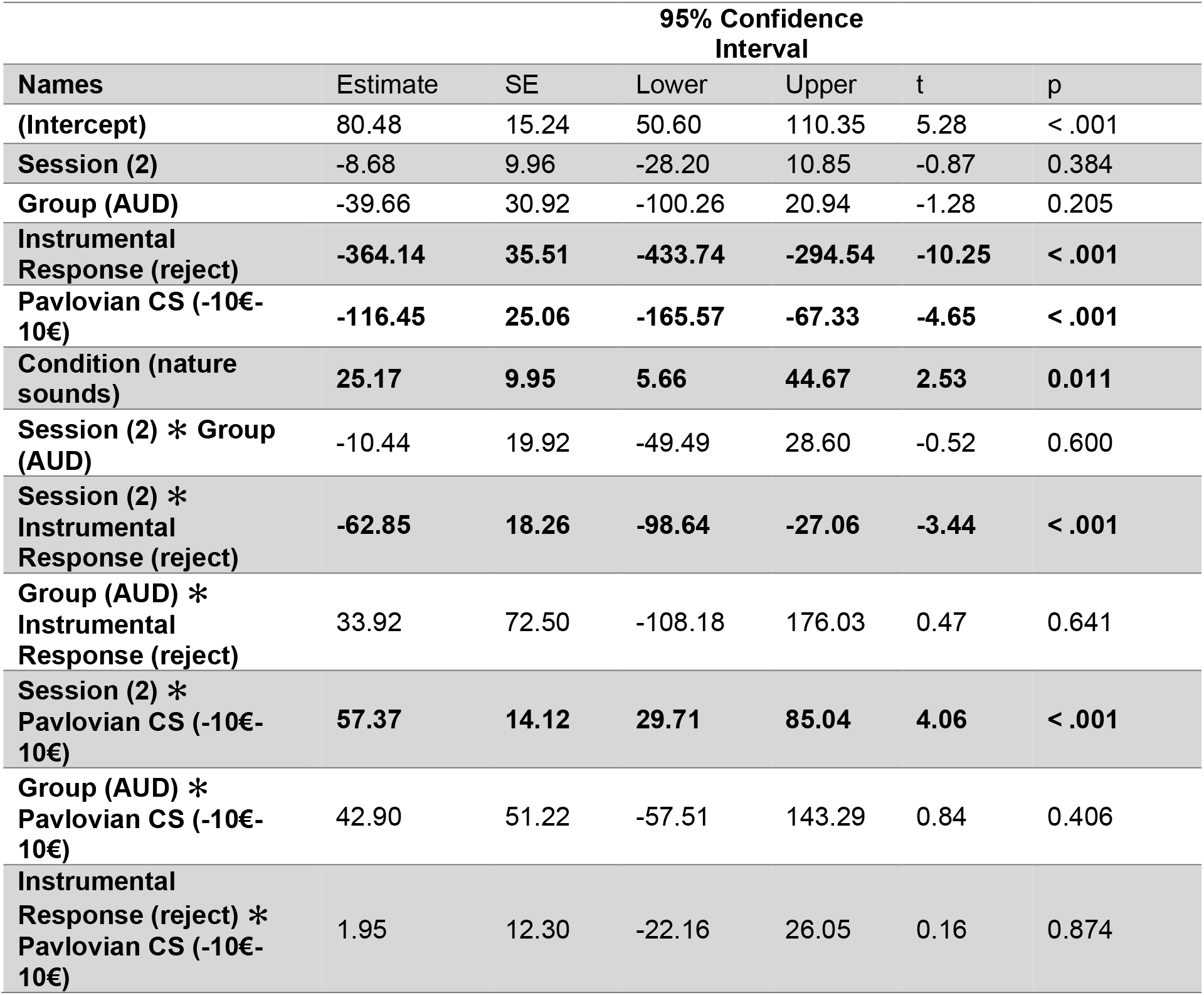
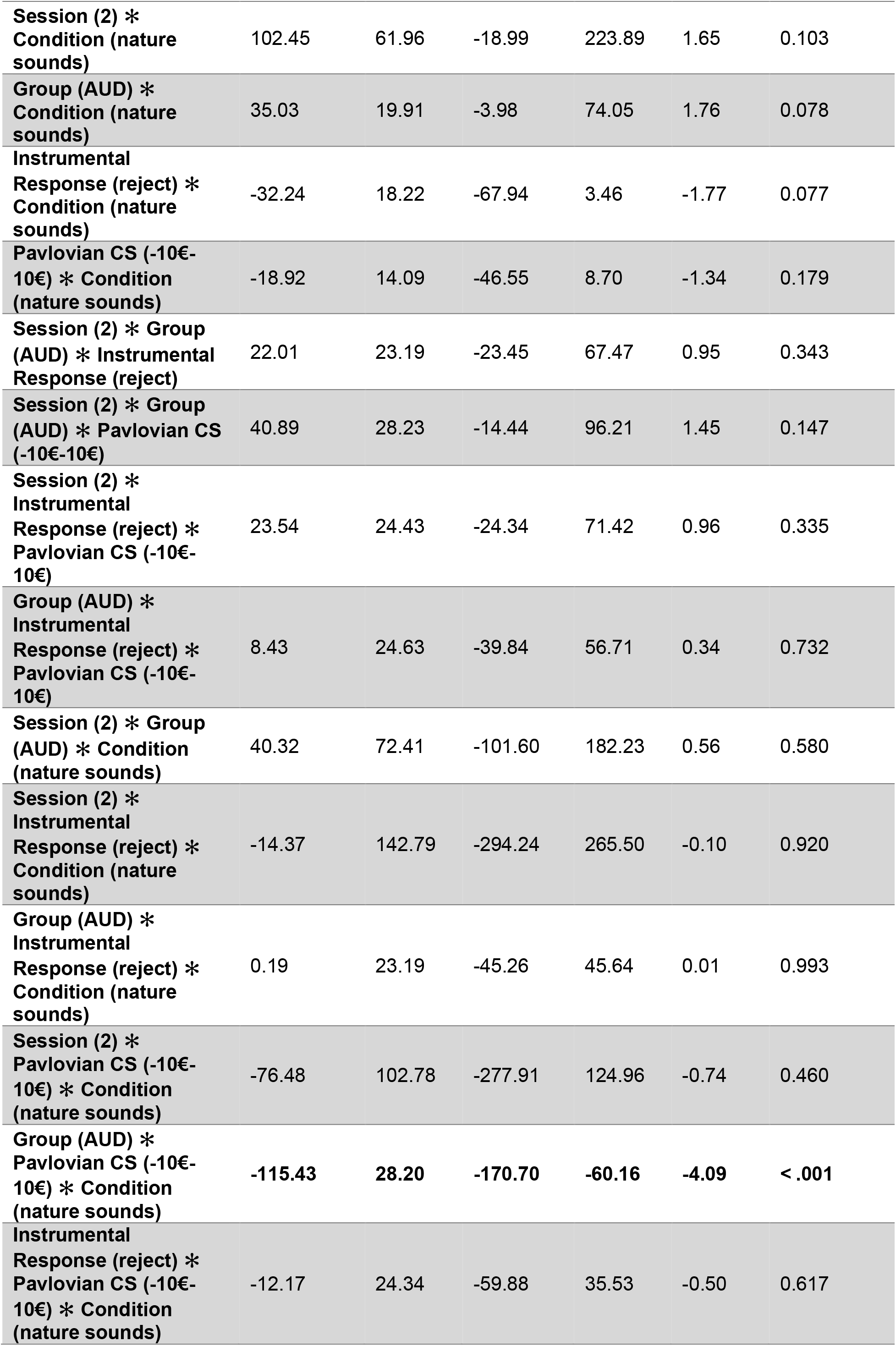
Fixed effects parameter estimates

In line with our hypotheses, there was an overall effect of Pavlovian CS on peak velocity (10€ -10€: estimate -116.45; t -4.65; p< .001), specifically, the positive Pavlovian CS was approached faster (mean = 80.49; SE = 15.2) than the neutral CS (mean = 6.02; SE = 11.8) while the negative CS was avoided (mean = -35.97; SE = 15.9). As expected, there was an effect of instrumental response (estimate -364.14; t -10.25; p< .001). Moreover, we found condition to be significant (estimate 25.17; t = 2.53; p =0.011), indicating a generally faster peak velocity after the nature sounds condition across all CS. Additionally, we found session to interact with instrumental response (estimate -62.85; t -3.44; p< .001), indicating a higher contrast between collect and reject trials (session 1 estimate -367, SE = 37.1; session 2 estimate -407, SE = 37.4) that implies better accuracy in the second session concerning instrumental contingencies. However, instrumental response did not interact with group (estimate 33.92; t 0.47; p=0.64) or condition (estimate -32.24; t -1.77; p=0.08) which suggests that these factors do not impact the instrumental performance in the transfer phase. Furthermore, we found a significant interaction between Pavlovian CS and session (estimate 57.37; t = 4.06, p< 0.001), here the PIT effect was less pronounced in the second session (please see table 4 for means and standard errors).

**Table 4.** Estimated marginal means

| Pavlovian CS | Session | Mean | SE | df | 95% Confidence Interval |  |
| --- | --- | --- | --- | --- | --- | --- |
|  |  |  |  |  | Lower | Upper |
| 10€ | 1 | 84.8 | 15.8 | 70.6 | 53.3 | 116.34 |
| 0 | 1 | -28.8 | 12.6 | 77.6 | -53.8 | -3.77 |
| -10€ | 1 | -60.3 | 16.4 | 70.4 | -93.1 | -27.52 |
| 10€ | 2 | 76.1 | 16.3 | 78.7 | 43.8 | 108.51 |
| 0 | 2 | 40.8 | 13.1 | 89.9 | 14.9 | 66.74 |
| -10€ | 2 | -11.6 | 16.9 | 77.6 | -45.2 | 21.95 |

We did not find an interaction effect of condition*Pavlovian CS (estimate -18.92, t = -1.34, p = 0.18), nor group*Pavlovian CS (estimate 42.90, t = 0.84, p= 0.41). Interestingly, however the interaction between group * condition * Pavlovian CS (10€ -10€: estimate -115.43; t -4.09; p < .001) indicates that after the body scan condition the PIT effect was reduced in the AUD group only (please see figure 3 for an illustration of this interaction effect).

**Figure 3.**
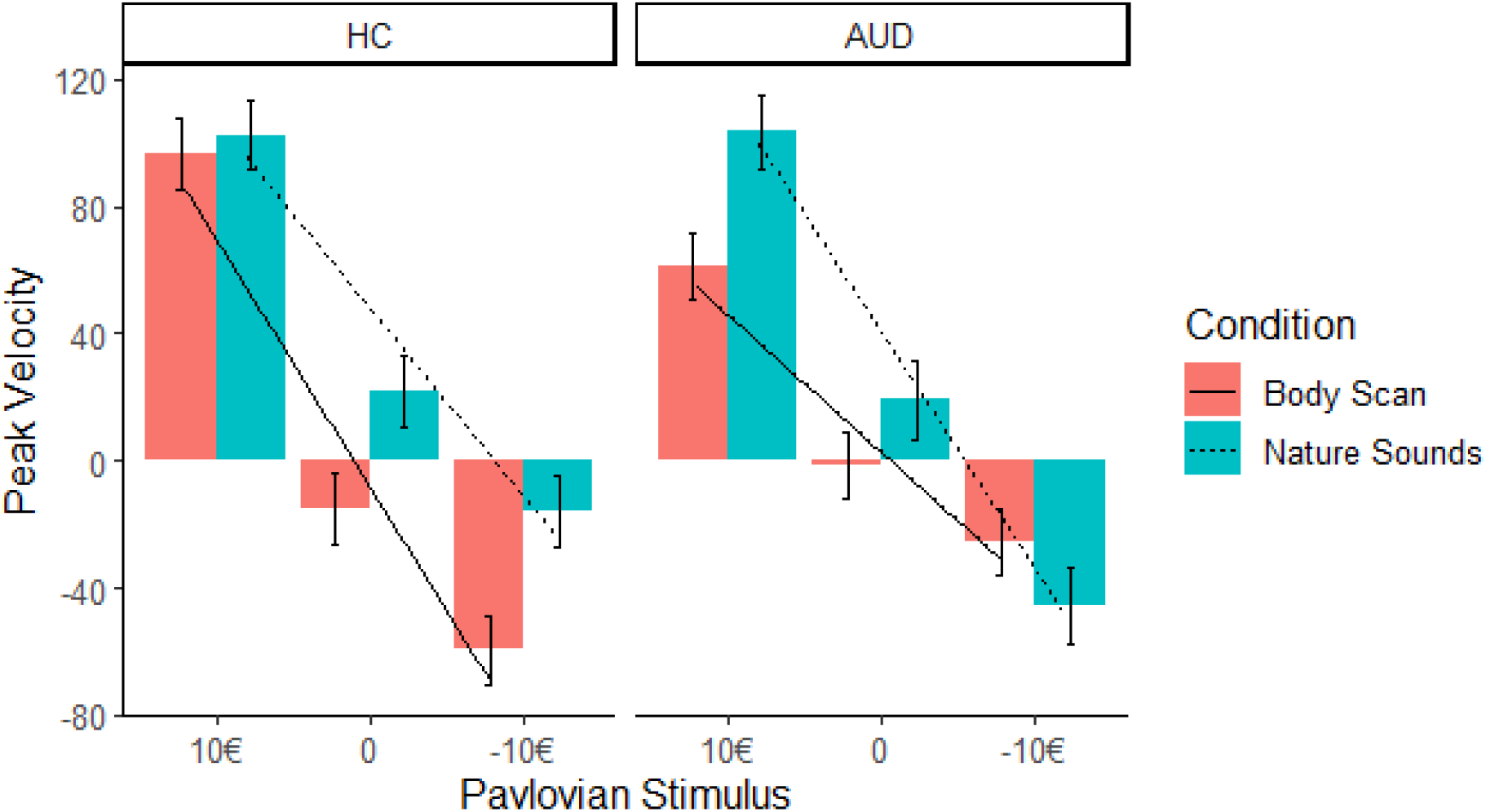
In AUD participants, the slope of peak velocity regressed on Pavlovian CS was not significant, indicating a decreased PIT effect after the body scan versus nature sounds condition in the AUD group only.

Simple effects analysis indicated that the effect of Pavlovian CS on peak velocity in the body scan condition was not significant in the AUD group (F = 1.19, p = 0.31). This contrast indicates the non-significance of the PIT effect in the AUD group after administration of the body scan. In turn, it was significant in HC across both conditions (body scan: F = 12.28, p < .001; nature sounds: F = 6.58, p = 0.002) as well as in the nature sounds condition in AUD (F = 5.42; p =0.006).

The results of the mixed ANOVA indicated no effect of the intervention conditions on change in self-reported vigilance (p=0.5), relaxation (p=0.51) and alertness (p=0.68). There was also no effect of group on the change in vigilance (p=0.95), relaxation (p=0.63) and alertness (p=0.30).

In addition, testing for an association among of trait mindfulness, stress and ADS, we found that perceived stress assessed by the PSS was negatively correlated with trait mindfulness assessed by FFMQ in both groups (AUD r = -0.7, p < 0.05; HC r = -0.51, p < 0.05). In AUD participants, PSS score positively correlated to ADS scores (r = -0.47, p < 0.05). (*Please see supplementary figures 1 and 2*). However, there was no association between ADS and FFMQ scores (r = -0.19, p > 0.05). Mediation analysis of the effect of FFMQ on ADS through PSS scores indicated a significant indirect effect (estimate = -.10; 95% CI = -.2169; -.0205), meaning that FFMQ is negatively associated with ADS though reduction of PSS scores.

## Discussion

To examine the underlying mechanisms of the efficacy of MBIs in addictive disorders, we assessed the influence of a brief mindfulness meditation on the magnitude of the PIT effect in AUD participants and healthy controls. Our results demonstrated a significant effect of the body scan intervention in reducing the PIT effect in the AUD cohort but not in healthy controls. Next to this, we could not find group differences in the slope of the PIT effect between our AUD and HC sample.

Using a newly developed PIT paradigm that utilizes a joystick response device, we detected a significant association between Pavlovian CSs and instrumental behavior in accordance with the related monetary reward. In contrast to neutral stimuli, the Pavlovian CS that were associated with monetary gain were approached faster. However, in contrast to findings by Belanger at al. (29), who observed a symmetrical effect of Pavlovian CS on approach and avoidance behavior, we found the negatively valanced Pavlovian CS to be avoided to a lesser extent than approach towards positive Pavlovian CS. Overall this finding corroborates the research on the influence of motivation on the magnitude of instrumental responding (39). The nature of the within-subjects design led to repeated administration of the PIT paradigm. While the order of the intervention conditions across sessions was randomized, we included session as a factor that indeed contributed to our model. In our sample, we found that the magnitude of the PIT effect decreased in the second session relative to the first one. Assessing effects of therapeutic interventions warrants outcome measures with good psychometric properties (40); here it is worth mentioning that versions of the PIT paradigm have been ascribed a low test-retest reliability (41). In light of this, the temporal stability of a variation of the PIT task used in this study has been previously investigated (42). Here, a paradigm that consisted of the same stimuli but utilized a button-box response device, was administered on two consecutive days. Analysis of intra-class correlation coefficients (ICC) for subject-specific PIT effects revealed a moderate temporal and moderate to good internal consistency (42). So, in summary, the proficient psychometric properties of our PIT task in combination with randomized intervention administration render our results well-founded and reliable.

Surprisingly, we did not find the AUD and HC group to differ in magnitude of the PIT effect when averaging across both intervention conditions. This stands in contrast to previous research that demonstrated an enhanced PIT effect in a sample of males at risk for AUD compared to low-risk consumers (43). In extension of these findings, Chen et al. (44) also found participants with risky drinking patterns to be more prone to be influenced by monetary Pavlovian CS compared to low-risk drinkers. These behavioral results were accompanied by functional neuroimaging findings that associated the strength of the behavioral PIT effect to decreased prefrontal activation as well as decreased prefrontal-limbic connectivity (44). In various studies that included patients with AUD and compared them with HC, an increased PIT effect was found as well (11). However, one of those studies found this group effect to be driven by the negative Pavlovian CS condition (11, 14). In addition, the magnitude of the PIT effect could distinguish abstainers from prospective relapsing patients (13). Sommer et al. (13) specifically observed that appropriate inhibitory behavior was disrupted by positive Pavlovian CS. These results stand in contrast to the lack of group differences between our AUD and HC samples, however, the employed response devices render the paradigms’ conditions dissimilar. In contrast to the button-box version used by the listed studies which requires the inhibition of a response, the joystick version demands an active avoidance of negative instrumental stimuli. Here, the active avoidance might not capture the dysfunctional inhibitory control that was previously found in prospectively relapsing AUD patients. Moreover, other studies also failed at detecting an increased PIT effect in populations with various substance use disorders (45-49). The mixed data situation has been proposed to result from discrepancies between the paradigms as well as systematic differences between AUD samples (42).

It has been suggested that prolonged drug exposure over the course of addiction, renders environmental cues to exert higher motivational impact on drug consumption by activating positive associations in cost of distinctive drug-memories (50). Thus, in contrast to the notion that the PIT effect might predispose for development of AUD, its’ magnitude might increase over the course of disease, providing an explanation why our subclinical sample might have not displayed a comparatively increased PIT effect.

Finally, in the AUD group, we found the body scan meditation to reduce the strength of the PIT effect. In fact, the effect of (negative) Pavlovian CS on instrumental behavior was rendered non-significant. In contrast, the HC group did not show any differences in PIT effect between conditions. This comparatively heightened susceptibility to the mindfulness intervention in the AUD group is difficult to interpret. Through a learning criterion in the instrumental learning phase, we ensured homogeneous accuracy during the transfer phase. Our results also showed no impact of group or condition on instrumental performance, which rules out the possibility of accuracy impacting the PIT effect. Additional analyses showed that the efficacy of the body scan intervention in AUD cannot be explained by subjective ratings of intervention-induced vigilance and alertness. As intended, relaxation was affected by both conditions equally. As the body scan is meant to induce a mindful state, we wanted to capture this state specifically and control for relaxation effects. We therefore employed a very strict nature sounds control condition instead of a waiting condition. However, subjective ratings on a visual analog scale might have not captured the effects as well as e.g. physiological measures such as heart rate variability (HRV). Past research has shown that mindfulness meditation induces changes in various measures of HRV (for a review, see 51). Furthermore, trait mindfulness did not differ between AUD and HC groups, ruling out a possible moderation effect of dispositional mindfulness on intervention effects that have been indicated by previous research (52). We could speculate that motivational aspects in the susceptibility to the mindfulness intervention might have played a role; in this case participants with AUD could have shown increased engagement during the mindfulness intervention compared to HC.

Furthermore, FFMQ-based trait mindfulness was negatively associated with perceived stress. Perceived stress in turn is positively correlated with dependence severity in AUD participants. Results of a mediation analysis indicated that mindfulness indirectly affects dependence severity through decreasing perceived stress. A body of research suggests a generally negative relationship between trait mindfulness and substance abuse and it has been suggested that this relationship is in part due to alterations in stress reactivity (53, 54). Synthesizing these findings with evidence for the role of stress as well as mindfulness in cue-reactivity processes, it is warranted that a thorough mindfulness training could reduce the effect of cues on maladaptive behavior.

Besides the described findings, our study has several limitations that we want to address. First, our groups contained unproportional gender distributions. The HC group is characterized by a high number of female subjects while the AUD sample contained mostly male participants. Concerning the AUD sample, our numbers reflect the epidemiological situation as men are diagnosed with AUD about four times as often as women (55). However, it has been repeatedly shown, that PIT effects are not affected by confounders such as age or gender (42). Another limitation is the comparatively small sample size that could have prevented us from detecting group differences in PIT effects as previous studies that noted positive results have used larger sample sizes (11-14, 43, 44). Finally, although we detected an effect on the magnitude of the PIT effect in AUD using an ultra-brief body scan meditation, the influence of mindfulness interventions on addiction-relevant cue reactivity should be investigated in the context of a comprehensive mindfulness training such as MBRP.

In conclusion, our study provides interesting results that show that Pavlovian CS exert effects on instrumental responding and that this effect can be decreased by application of a brief mindfulness intervention. This reduction in magnitude of the PIT effect was seen in AUD participants only. In sum, our results contribute to the understanding of the underlying mechanisms of MBIs in substance use disorders and suggest that mindfulness training could be a suitable intervention strategy to reduce the impact of environmental cues on maladaptive behavior in the context of AUD.

## Data Availability

All data produced in the present study are available upon reasonable request to the authors.

