## Supplementary Material for "Effects of a Brief Mindfulness-Based Intervention on Pavlovian-To-Instrumental Transfer in Alcohol Use Disorder"

Mediation analysis

I order to test for a conditional effect of trait mindfulness (FFMQ) on dependence severity (ADS) through perceived stress (PSS), a mediation model was constructed. FFMQ score was the predictor variable, with PSS score as the mediator. The outcome variable was ADS score. The hypothesized mediation model was tested via bootstrapping approach using the “PROCESS" macro, model 4, version 4.2 (Hayes, 2017) in SPSS version 28 (IBM SPSS Statistics for Windows; Armonk, NY: IBM Corp) with bias-corrected 95% confidence intervals (n = 5000). Significant effects are supported by the absence of zero within the confidence intervals.

Query trials

|  |  |  | 95% Confidence Interval | |  |  |
| --- | --- | --- | --- | --- | --- | --- |
| Names | Estimate | SE | Lower | Upper | t | p |
| (Intercept) | 6.74 | 0.308 | 6.133 | 7.342 | 21.86 | < .001 |
| condition (NS) | 0.01 | 0.406 | -0.786 | 0.806 | 0.02 | 0.98 |
| session (1) | 0.09 | 0.406 | -0.702 | 0.889 | 0.23 | 0.82 |
| condition (NS) ✻ session (1) | -1.72 | 1.233 | -4.133 | 0.702 | -1.39 | 0.17 |

### Supplementary figure 1. Condition and session effects on performance in forced choice task

| Fixed Effect Omnibus tests |  |  |
| --- | --- | --- |
|  | F | p |
| **Session** | **39.90367** | **< .001** |
| Group | 0.38722 | 0.536 |
| **Instrumental Response** | **96.57844** | **< .001** |
| **Condition** | **6.98312** | **0.008** |
| **Pavlovian Stimulus** | **13.89861** | **< .001** |
| Session ✻ Group | 3.71789 | 0.054 |
| **Session ✻ Instrumental Response** | **18.66953** | **< .001** |
| Group ✻ Instrumental Response | 0.54521 | 0.463 |
| Session ✻ Condition | 1.64389 | 0.205 |
| **Group ✻ Condition** | **8.53802** | **0.003** |
| **Instrumental Response ✻ Condition** | **8.37615** | **0.004** |
| **Session ✻ Pavlovian Stimulus** | **16.26291** | **< .001** |
| Group ✻ Pavlovian Stimulus | 0.39418 | 0.676 |
| Instrumental Response ✻ Pavlovian Stimulus | 1.12088 | 0.326 |
| Condition ✻ Pavlovian Stimulus | 0.73954 | 0.477 |
| Session ✻ Group ✻ Instrumental Response | 0.85608 | 0.355 |
| Session ✻ Group ✻ Condition | 0.82548 | 0.367 |
| Session ✻ Instrumental Response ✻ Condition | 0.03333 | 0.856 |
| Group ✻ Instrumental Response ✻ Condition | 0.00161 | 0.968 |
| Session ✻ Group ✻ Pavlovian Stimulus | 2.01492 | 0.133 |
| Session ✻ Instrumental Response ✻ Pavlovian Stimulus | 0.69771 | 0.498 |
| Group ✻ Instrumental Response ✻ Pavlovian Stimulus | 1.06870 | 0.343 |
| Session ✻ Condition ✻ Pavlovian Stimulus | 0.63501 | 0.533 |
| **Group ✻ Condition ✻ Pavlovian Stimulus** | **8.71862** | **< .001** |
| Instrumental Response ✻ Condition ✻ Pavlovian Stimulus | 0.37238 | 0.689 |
| Session ✻ Group ✻ Instrumental Response ✻ Condition | 0.12563 | 0.724 |
| Session ✻ Group ✻ Instrumental Response ✻ Pavlovian Stimulus | 0.91533 | 0.400 |
| Session ✻ Group ✻ Condition ✻ Pavlovian Stimulus | 2.82753 | 0.067 |
| Session ✻ Instrumental Response ✻ Condition ✻ Pavlovian Stimulus | 0.05554 | 0.946 |
| Group ✻ Instrumental Response ✻ Condition ✻ Pavlovian Stimulus | 0.64716 | 0.524 |
| Session ✻ Group ✻ Instrumental Response ✻ Condition ✻ Pavlovian Stimulus | 0.44144 | 0.643 |

Supplementary table 3. Results of the linear mixed analysis type III test of fixed effects

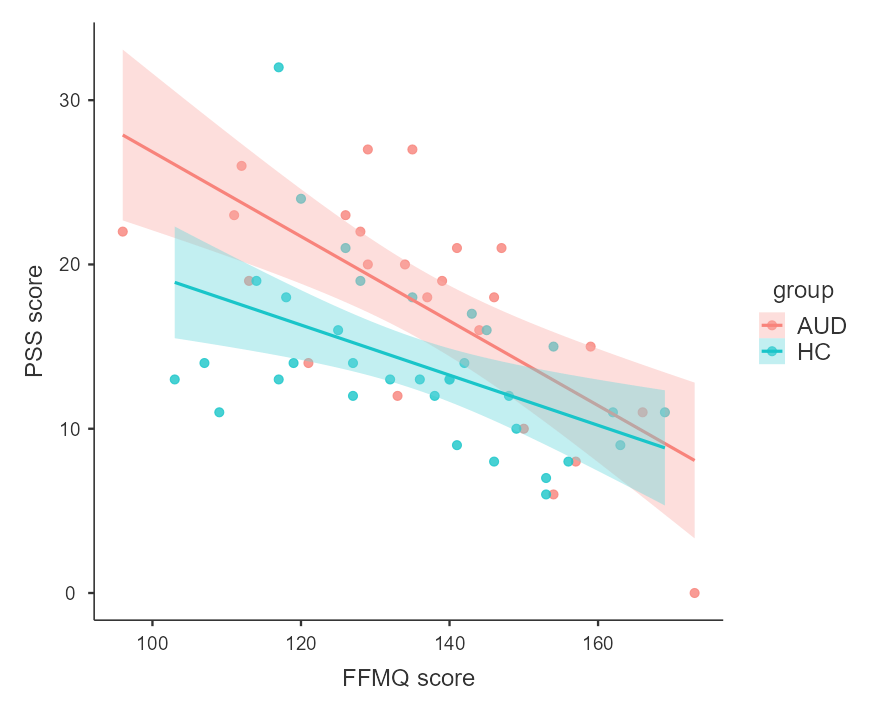

Supplementary figure 1. Scatterplot with regression lines to illustrate the relationship between perceived stress (PSS) and trait mindfulness (FFMQ). Blue reflects data from healthy controls and red reflects data from alcohol use disorder participants.

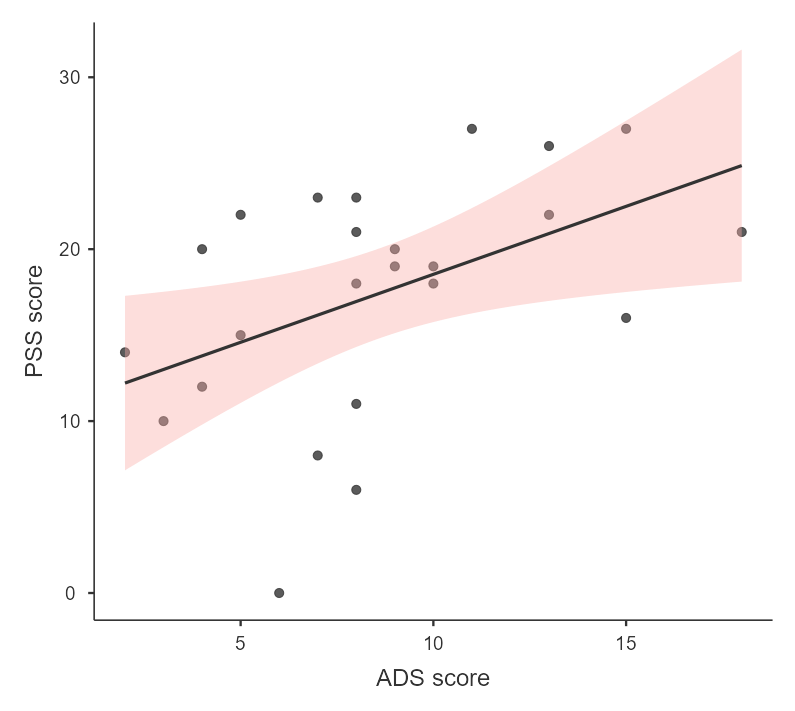

Supplementary figure 2. Scatterplot with a regression line to illustrate the relationship between perceived stress (PSS) and dependence severity (ADS) in participants with alcohol use disorder.
